# DLBCLone: A unified framework for neighbourhood-based genetic subtyping of lymphomas

**DOI:** 10.1101/2025.09.18.25335809

**Authors:** Luke Klossok, Kostiantyn Dreval, Manuela Cruz, Jasper C.H. Wong, Sierra Gillis, Brett Collinge, Christian Steidl, David W. Scott, Laura K. Hilton, Ryan D. Morin

**Author notes:** **Corresponding Author Dr. Ryan D. Morin** Department of Molecular Biology and Biochemistry Simon Fraser University 8888 University Drive Burnaby, BC, Canada.

## Abstract

Genetic subtyping of diffuse large B-cell lymphoma (DLBCL) has been slow to gain clinical adoption. Available classifiers either leave many tumours unclassified or depend on exome-wide features and copy-number profiles, which are not always available in routine practice. We introduce DLBCLone, a neighbourhood-based framework that enables panel-aware genetic subtyping compatible with existing taxonomies. DLBCLone learns a 2-D reference map of mutation profiles (UMAP) from a labeled training cohort, freezes this map, and deterministically projects new cases into the same latent space. Class labels are then inferred by weighted K-nearest neighbours, limiting over-assignment by considering the local density of unclassified neighbours. By default, classification thresholds optimize per-class balanced accuracy, but can be adjusted to suit study needs. The framework is intended to emulate (or “clone”) existing schemas such as LymphGen or DLBClass.

Trained on a harmonized cohort of 2,130 DLBCLs, DLBCLone classifiers for different gene panels achieved consistently improve classification rates relative to fixed-threshold baselines while maintaining a reasonable per-class performance. On an in-house cohort of 323 patients, it assigned an additional 98 samples without compromising accuracy relative to LymphGen. On an external exome-sequenced subset from a 1,001-patient cohort, DLBCLone achieved a 51% classification rate (vs 36% for LymphGen) at an overall accuracy of 0.70. Compared with another LymphGen approximator (LymphPlex), DLBCLone reached a 74% classification rate (vs 55%). In general, the DLBCLone-reclassified tumours had molecular features consistent with their new labels.

DLBCLone provides a deterministic, reproducible, and extensible approach to genetic subtyping under real-world constraints, facilitating prospective studies that rely on either targeted panels or more comprehensive sequencing strategies. DLBCLone is open source and available in the GAMBLR.predict package (https://github.com/morinlab/gamblr.predict).

## Introduction

Diffuse large B-cell lymphoma (DLBCL) comprises biologically diverse cancers unified by morphology. Early genomic work divided DLBCL into two gene expression-based molecular subgroups with prognostic implications, with ABC-DLBCL generally showing worse outcomes than GCB-DLBCL^1^. More recently, a “dark-zone” gene expression signature (DZsig) was proposed to further sub-divide GCB-DLBCL, with DZsig-positive cases having outcomes comparable to ABC-DLBCL^2^.

Comprehensive sequencing studies have identified >125 recurrently mutated genes in DLBCL, revealing patterns of co-occurrence and mutual exclusivity that motivated genetically defined subgroups^3,4^. This work culminated in LymphGen, a probabilistic classifier that assigns tumors to EZB, MCD, BN2, ST2, N1, and A53^5^. EZB can be further sub-divided based on presence of the DZsig gene expression signature or, in lieu of this, MYC translocation status. A separate group proposed a five-class system with each group (clusters C1–C5) broadly aligning with a LymphGen counterpart, with the exception of N1. Results from a third study indicated that at least one of these classes (ST2) could be further sub-divided. Differences across studies reflect both methodological choices and feature scope: for example, the panel-based analysis by Lacy *et al* could not recover A53/C2 (largely driven by copy-number variants (CNVs)) and did not reproduce N1. A later update from this group reassigned truncating *NOTCH1* cases accordingly, bringing the “HMRN” system closer to LymphGen^6–8^.

As genetic subgroups gained traction, their clinical relevance—and potential therapeutic vulnerabilities—have been explored, including some encouraging post hoc analyses supporting rational combinations in specific subgroups^9^. Despite this, adoption has been slow while the community evaluates technical and analytical barriers to bringing genetic classification to real-world settings. The deliberately conservative thresholds used by LymphGen leave ∼39–47% of cases unclassified (“Other”), which complicates trial accrual^5^. While the original description of the C1-C5 system suggested a nearly complete classification rate, these groupings were originally assigned by clustering, which cannot be used to assign new samples due to the inter-dependence of all samples on cluster membership. A reproducible tool for this (DLBClass) became available only recently, leaving no options other than LymphGen in the interim^10^. In this setting, a demand for alternatives led to the emergence of simplified proxies for genetic classification^11^. LymphPlex, for example, uses a 37-gene panel to produce LymphGen-like labels (e.g., N1-like, ST2-like) and reports higher classification rates with modest panels^12^. To date, LymphPlex is the only framework that has been formally incorporated into a subtype-guided prospective trial^13,14^.

Despite the appeal of comprehensive sequencing, gene panels persist in clinical workflows, and yet no commercial panels have been designed around LymphGen/DLBClass features. Moreover, because current classifiers were optimized using the full feature universe, their performance can degrade unpredictably when specific genes or CNVs are missing. Collectively, these realities motivate the existence of a tool with flexibility for features and tuneable classification rates.

We present DLBCLone, which lets users examine how feature combinations map onto DL-BCL subtypes and build reproducible, panel-aware classifiers à la carte. Unlike fixed-schema methods, DLBCLone is system-agnostic. Given a sufficiently sized labeled (training) cohort, previously analyzed with an available classifier (e.g. LymphGen), DLBCLone can emulate genetic taxonomies and classify new cases deterministically. Rather than requiring the experimental data to cater to the requirements of the classifier, DLBCLone is designed to adapt to the available data while revealing their impact on overall and per-class accuracy.

We demonstrate the flexibility of this approach across a variety of gene panels by training on over 2000 DLBCLs from our harmonized cohort. We validate generalizability on independent, previously unseen, real panel-based datasets. Across these external cohorts, DLBCLone yields consistently higher classification rates than fixed-threshold baselines (e.g., LymphGen) while maintaining comparable or improved per-class balanced accuracy, supporting its suitability for prospective studies that rely on panels or exomes.

## Results

### Genetic features associated with LymphGen classes

Our training data comprises samples from numerous publications that were sequenced with either exome or whole genome sequencing (WGS) and accompanying *BCL2* and *BCL6* rearrangement status (**Methods**). Noting the range of sequencing depth and a strong association between low coverage and LymphGen classification rate, we began by eliminating samples with exceedingly poor coverage (**Figure 1**)^15^. The remaining 2130 samples with unambiguous LymphGen assignments were used for all subsequent analyses.

**Figure 1:**
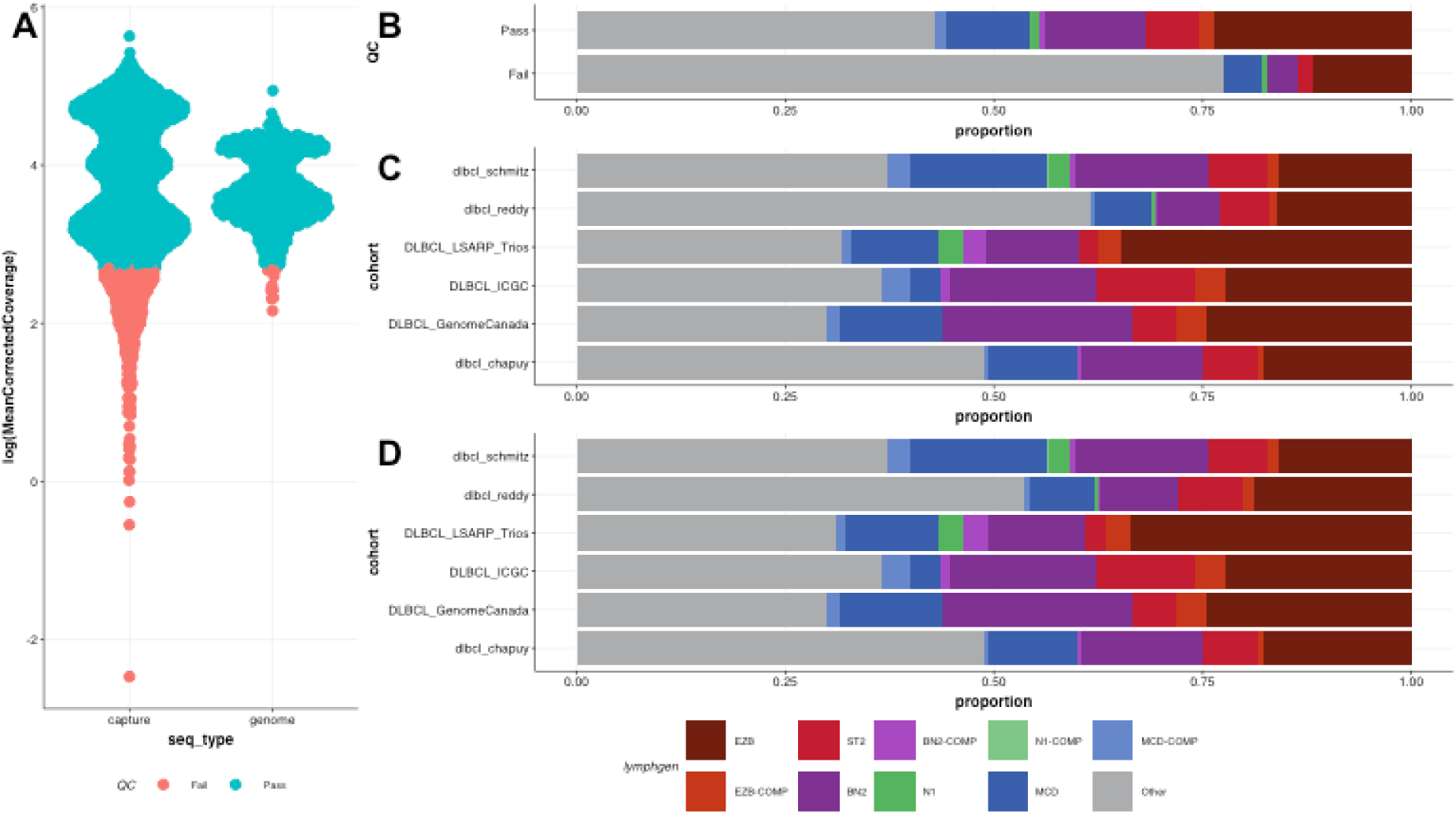
Association between sequencing depth and study on LymphGen class representation. **A)** The distribution of coverage values across all exome capture (left) and WGS (right) samples. The variability across the exomes was particularly striking, with more than 300 samples having less than 15x average corrected coverage. For the purposes of this analysis, we considered those as QC-Fail. **B)** There is a clear difference in the success of LymphGen on the samples that passed QC (top) compared to the remaining samples. The proportion of cases assigned to a non-Other class was 45% in the QC-Pass samples. Although the rate of most LymphGen classes was similar, N1 samples were not found in three of the largest cohorts. **C)** Considering the six largest studies with samples in this analysis, the Reddy cohort was the major contributor to a higher rate of LymphGen-Other. **D)** The distribution of LymphGen classes in the same cohorts after removing QC-Fail samples.

To aid in feature selection for DLBCL classification, we compared the frequency of genetic features between samples based on their assigned LymphGen class (**Methods**). Aiming to develop a flexible method suitable for either gene panels or comprehensive sequencing data, we focused on simple somatic mutations and translocations that are detected in routine clinical practice (*BCL2* and *BCL6*). For select genes, features were derived only from specific mutation classes. For example, we tabulated *NFKBIZ* 3^′^ UTR mutations and separately annotated *MYD88* hotspot and non-hotspot mutations. This analysis recovered nearly all the features used by LymphGen (**Supplemental Table S1**). Interestingly, 28 additional associations between mutations and at least one LymphGen class were identified (**Table 1**). While some of these associations were reported in the original description of these classes (e.g. *STAT6* and *TMEM30A*)^4^, their mutations are not utilized within LymphGen. Some of these genes are, however, utilized by the DLBClass tool, such as *TMSB4X*, *KLHL6*, *BRAF*, *STAT6*, *GNA13* and *TOX*^10^. For some of the remaining genes, the low incidence could explain why these associations were not previously detected. For example, *FCGR2B* mutations were in less than 4% of EZB samples and *CDKN2A* mutations occur in less than 7% of MCDs. Though mutations in these genes are not common, CNVs affecting thes loci have been described in GCB and ABC DLBCL, respectively^16–18^. When CNV features are given to LymphGen, the presence of *CDKN2A* deletions (but not mutations) contributes to MCD classification. Two novel features that were strongly enriched in BN2 are non-hotspot mutations in *MYD88* and *NFKBIZ* 3 UTR mutations. Although detectable by exome sequencing, coverage of these regions varies across lymphoma gene panels. As LymphGen does not use these features, we evaluated whether adding them to training improves classification accuracy and robustness.

**Table 1:** Novel features enriched per LymphGen class.

| Feature | Class | In-class incidence | Out-of-class incidence | P | Odds Ratio |
| --- | --- | --- | --- | --- | --- |
| FAS | BN2 | 0.167 | 0.085 | 0.0002930 | 2.150 |
| NFKBIE | BN2 | 0.078 | 0.031 | 0.0020045 | 2.657 |
| NFKBIZ 3' UTR | BN2 | 0.206 | 0.022 | 0.0000000 | 11.710 |
| TMEM30A | BN2 | 0.148 | 0.030 | 0.0000000 | 5.674 |
| TMSB4X | BN2 | 0.280 | 0.186 | 0.0015634 | 1.706 |
| TP73 | BN2 | 0.054 | 0.021 | 0.0083328 | 2.750 |
| BTK | EZB | 0.093 | 0.032 | 0.0000199 | 3.127 |
| FCGR2B | EZB | 0.042 | 0.000 | 0.0000000 | Inf |
| FOXO1 | EZB | 0.119 | 0.056 | 0.0001474 | 2.290 |

Table 1: Novel features enriched per LymphGen class.
| Feature | Class | In-class<br>incidence | Out-of-class<br>incidence | P | Odds Ratio |
| --- | --- | --- | --- | --- | --- |
| GNA13 | EZB | 0.202 | 0.098 | 0.0000011 | 2.317 |
| S1PR2 | EZB | 0.085 | 0.014 | 0.0000000 | 6.414 |
| STAT6 | EZB | 0.129 | 0.035 | 0.0000000 | 4.078 |
| TP53 | EZB | 0.315 | 0.162 | 0.0000000 | 2.377 |
| CD19 | MCD | 0.081 | 0.017 | 0.0000146 | 4.963 |
| CDKN2A | MCD | 0.071 | 0.030 | 0.0086631 | 2.447 |
| HNRNPD | MCD | 0.052 | 0.016 | 0.0040064 | 3.328 |
| IRF4 | MCD | 0.128 | 0.036 | 0.0000012 | 3.955 |
| PRDM15 | MCD | 0.085 | 0.039 | 0.0069268 | 2.298 |
| TMSB4X | MCD | 0.327 | 0.180 | 0.0000051 | 2.217 |
| TOX | MCD | 0.123 | 0.048 | 0.0001779 | 2.807 |
| BIRC3 | ST2 | 0.065 | 0.013 | 0.0005574 | 5.268 |
| BRAF | ST2 | 0.116 | 0.019 | 0.0000005 | 6.732 |
| KLHL6 | ST2 | 0.188 | 0.072 | 0.0000362 | 2.978 |
| NFKBIE | ST2 | 0.101 | 0.033 | 0.0007819 | 3.293 |
| PRKDC | ST2 | 0.109 | 0.050 | 0.0099676 | 2.307 |
| PTPN1 | ST2 | 0.043 | 0.008 | 0.0038031 | 5.605 |
| TBCC | ST2 | 0.080 | 0.012 | 0.0000218 | 7.088 |
| XBP1 | ST2 | 0.065 | 0.021 | 0.0067721 | 3.237 |

### Separation of DLBCL with different feature sets

To visualize patterns across many genetic features, we use the Uniform Manifold Approximation and Projection (UMAP), which reduces high-dimensional data into a two-dimensional map where nearby points have more similar mutation profiles. We began by exploring the relationship between selected gene sets and tumours assigned to different LymphGen classes. Across different gene sets, samples of the same class tended to occupy similar regions in UMAP space (**Figure 2**). The main exception was N1, defined solely by *NOTCH1* mutations. Our inhouse panel (LySeqST) showed separation comparable to the LymphGen feature set(**Figure 2C**)^19^. This was encouraging because this panel was developed, in part, to apply LymphGen to clinical samples. We noted varying degrees of overlap between ST2 and BN2, most pronounced with the LymphPlex panel and also seen with the Lacy panel (**Figure 2D,E**). The samples labeled Other by LymphGen were diffusely distributed with many lying in the proximity of *bona fide* class neighborhoods.

**Figure 2:**
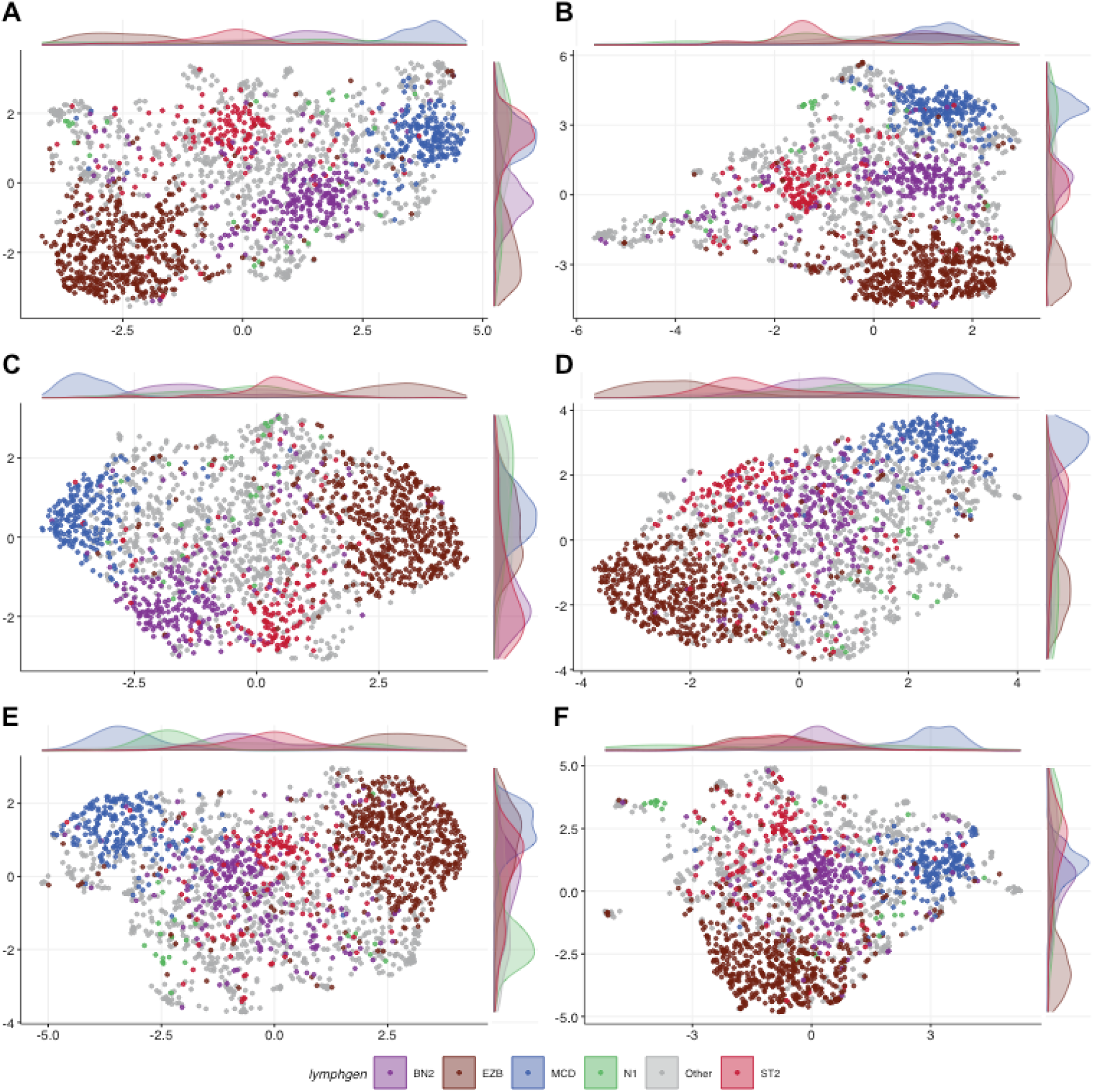
UMAP transformation of DLBCLs using different feature sets. Each plot shows the result of UMAP using the a subset of genes as described. *BCL2* and *BCL6* translocation status was used for A, B and D to be consistent with the methods used in the corresponding studies. All DLBCL samples assigned to a single class by LymphGen were included. Points are coloured based on their class as defined by LymphGen. **A)** All features that were significantly associated with a LymphGen class in our post-hoc analysis. **B)** The full set of features used by the LymphGen algorithm. **C)** The LySeqST 125-gene panel. **D)** A 93-gene panel used in the validation of LymphGen. **E)** 78 genes from Lacy et al. This includes only the genes that were used in their final clustering, further restricted to the genes considered Tier 1 DLBCL genes.^3^ **F)** The 37 genes used in the LymphPlex algorithm.

### DLBCLone performance without tuning

We reasoned that if a DLBCL sample could be mapped to this latent space, the class of previously analyzed (i.e. training) samples in the same local neighborhood could allow inference of its LymphGen class by proximity. Considering that some Lymphgen-Other samples have mutations in genes associated with one of the LymphGen classes, we speculated that a datadriven neighbor-based approach could achieve a classification rate exceeding LymphGen by tuning the threshold between assigning a sample to a class rather than Other. To support different use cases, our approach assigns up to two classifications to each sample. The first represents a more generous (“greedy”) strategy that attempts to maximize classification rate. The second results from an optimization between classification rate and accuracy, effectively increasing the barrier for samples to be assigned to any class.

To illustrate the general approach, we implemented a DLBCLone model to approximate LymphGen utilizing all genetic features we determined to be significantly associated with a class. The greedy assignments exceeded ∼74% accuracy for all but one class. Each of MCD and EZB were classified with ∼95% accuracy. Unfortunately, a minority of the N1 samples were correctly assigned (**Figure 3A**), which was not surprising given the lack of any clear cohesion of these samples in the UMAP. The outgroup-optimized classifications reduced the classification rate from 96% to 78.3% with only modest reductions in accuracy (**Figure 3B**).

**Figure 3:**
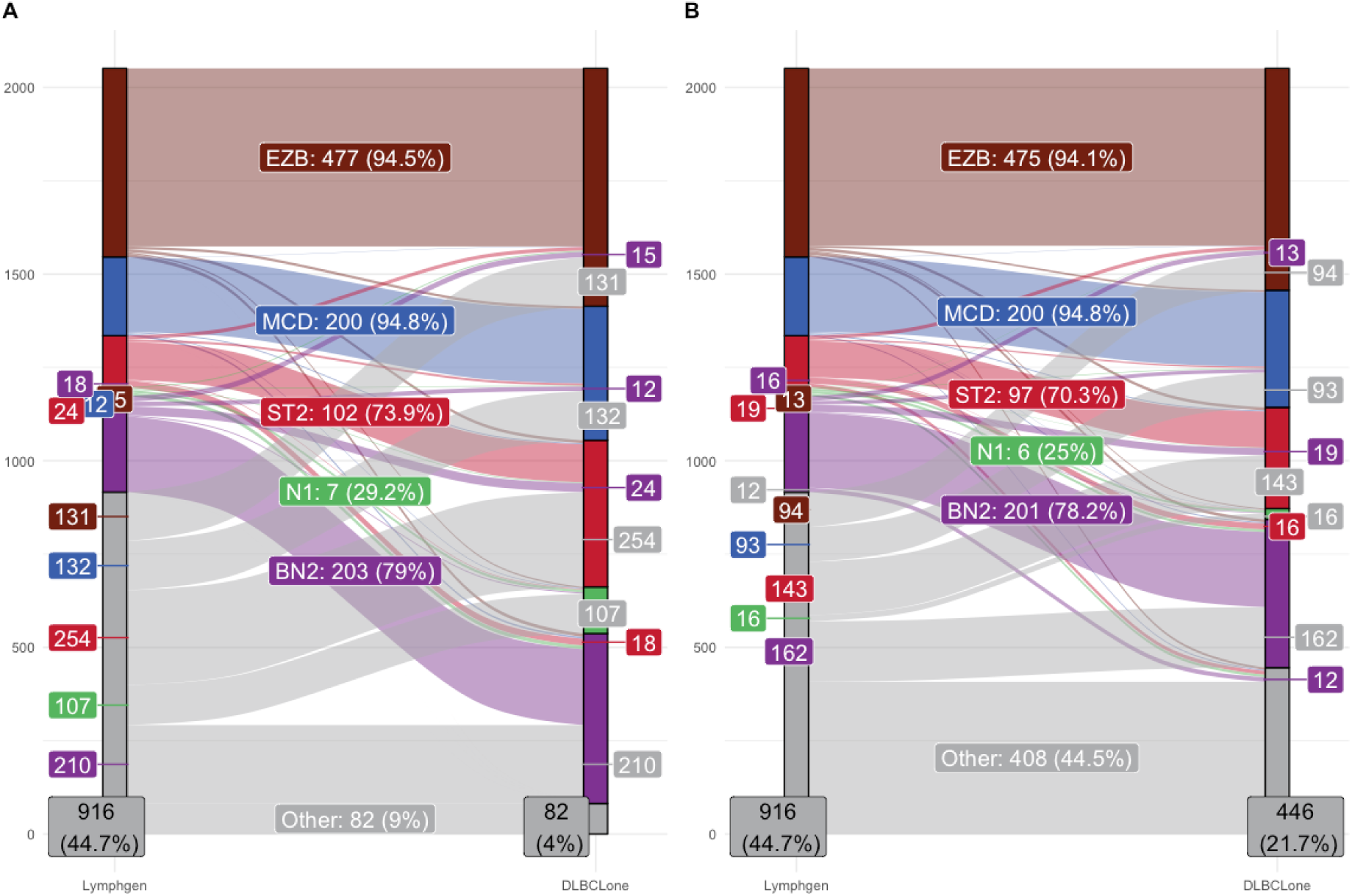
Accuracy of DLBCLone greedy and optimized labels. A basic DLBCLone classifier was trained using all features significantly associated with a LymphGen class (Supplemental Table S1). Alluvial summaries of the relationship between LymphGen assignments and the LymphGen class assigned by DLBCLone. Each ribbon is coloured according to the original (“true”) class of the training sample. The total number and percentage of training samples assigned to the same class by DLBCLone is indicated in boxes in the centre. The coloured boxes on either side indicate the number of samples that changed to another class. Outer boxes are repeated on both sides for clarity and coloured based on the opposite class. **A)** DLBCLone without optimization (greedy) **B)** DLBCLone optimized for balance between classification and Other.

### Influence of feature repertoire and weighting

While this performance was encouraged, we explored strategies to further improve accuracy, particularly for under-performing classes such as N1. To bolster the technique, we next investigated approaches to enhance this separation, using the vizualizations of UMAP transformations as a guide. We were particularly motivated to improve separation of classes with more diffuse patterns in the UMAP such as N1 and to reduce the overlap between BN2 and ST2. The weighting strategy we adopted combines sets of related features that support a given class into meta-features, a technique also used by DLBClass. To maintain flexibility, we do not perscribe which genes should contribute to the meta-features. In some scenarios, a user may prefer to optimize recovery of only certain classes, in which case they may choose to provide meta-features only for those. For the following analyses, we used a set of meta-features for each of the LymphGen classes (**Table 2**).

**Table 2:** Mutations used to construct meta-features.

| Meta-feature | Genes |
| --- | --- |
| ST2-meta | *SGK1, DUSP2, TET2, SOCS1* |
| N1-meta | *NOTCH1* |
| EZB-meta | *EZH2, BCL2* SV |
| MCD-meta | *MYD88<sup>L265P</sup>, CD79B, PIM1* |
| BN2-meta | *BCL6* SV, *NOTCH2, SPEN, CD70* |

We sought to quantify the influence of different feature combinations on the ability to infer genetic subroup of a sample based solely on the LymphGen class of samples in its neighborhood. Importantly, the LySeqST panel was designed to cover all LymphGen features whereas the Lacy and LymphPlex panels are each incomplete. We created UMAP transformations for the three panels shown in **Figure 3** using the features present in each panel and the metafeatures described above. **Figure 4** shows the original UMAPs for the three feature sets using meta-features (**A–C**). There is a more clear separation of most N1 samples from the remaining class neighborhoods. The ST2 and BN2 neighborhoods also show a greater degree of separation, particularly in the UMAP based on our LySeqST panel. The effect of freezing the reference map and projecting the same samples with the locked transform can be seen as well (**Figure 4D–F**). Although the points shift slightly, they preserve their local neighbourhood structures. This deterministic projection is essential for prospective use: new cases can be added and classified in the same coordinate system at any time without refitting or moving the reference map, ensuring consistent classification even when some data are unavailable—or intentionally held out—at training time. Finally, **Figure 4G–I** colours the same projected points by their new class assignment, illustrating how the local neighbourhoods correspond to common DLBCLone predictions.

**Figure 4:**
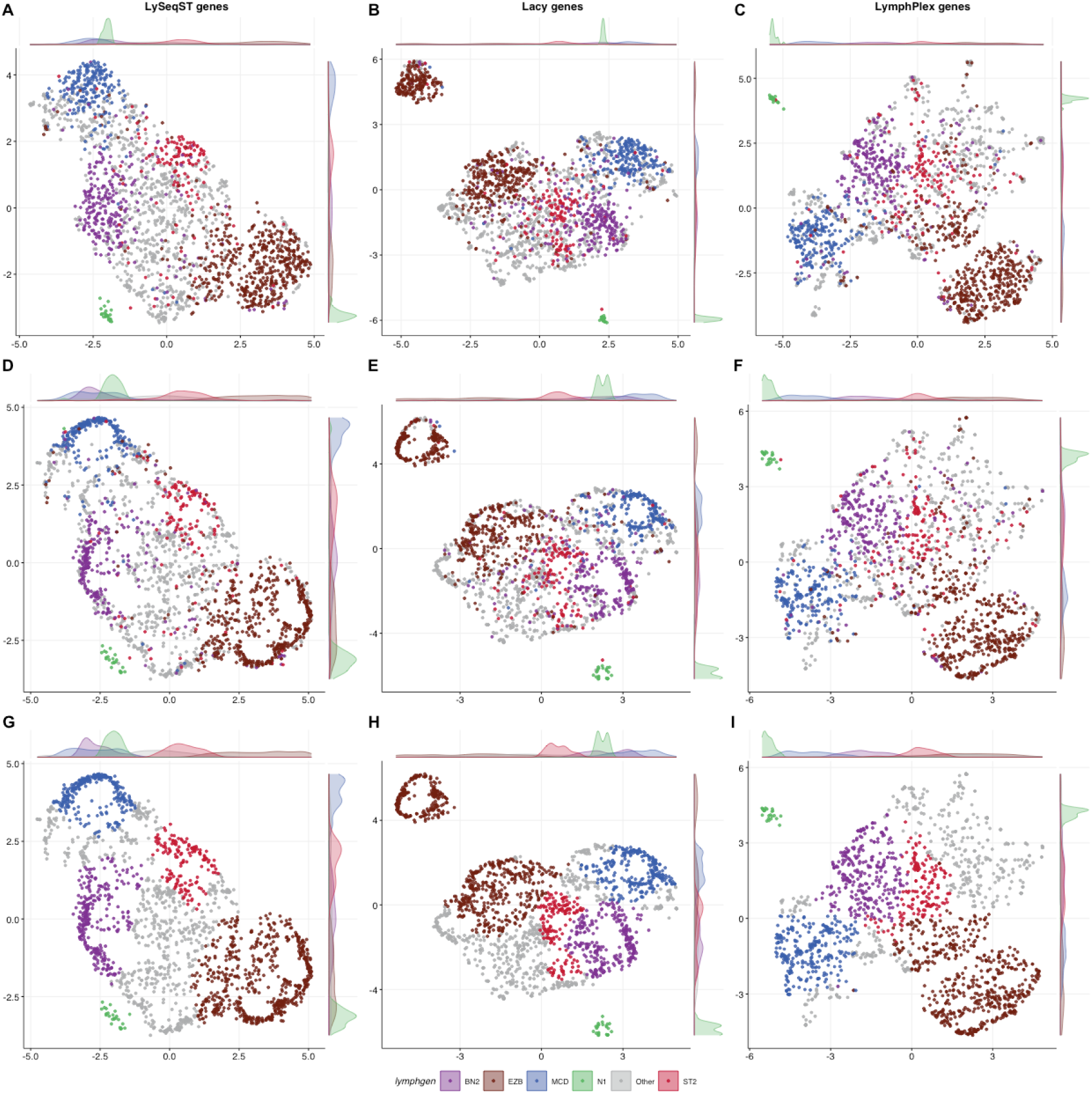
Latent representations with feature weighting. **A-C)** The initial UMAP transformation of the training samples based on the LySeqST genes (A), Lacy genes (B), or LymphPlex genes (C) with weighting applied to a core set of features for each class. Points are coloured by the LymphGen class of the sample. **D-F)** show the corresponding result after each sample is transformed using the stable UMAP model with umap_transform. **G-I)** The same points from D-F coloured by their predicted LymphGen class from the DLBCLone optimized model derived from this UMAP.

Overall, the classifiers derived from the three panels showed encouraging results (**Figure 5**). The DLBCLone-LySeqST classifier yielded the highest agreement across LymphGen groups, ranging from 61.6% (ST2) to 95.8% (N1), using the optimized predictions. With a classification rate of 65%, DLBCLone-LySeqST assigned 173 of the LymphGen-Other samples to a new class. The two smaller panels each achieved higher classification rates along with a slight reduction in accuracy across most classes (**Figure 5,B-C**). For DLBCLone-LymphPlex, based on the smallest panel, the accuracy per class ranged from 50% (for ST2) to 100% (for N1). One might conclude from this result that a smaller panel is preferrable for applications that require a higher classfication rate. However, we caution against this interpretation because the quality of classifications for the newly classified samples is difficult to assess, since they (by definition) conflict with what has been considered ground truth.

**Figure 5:**
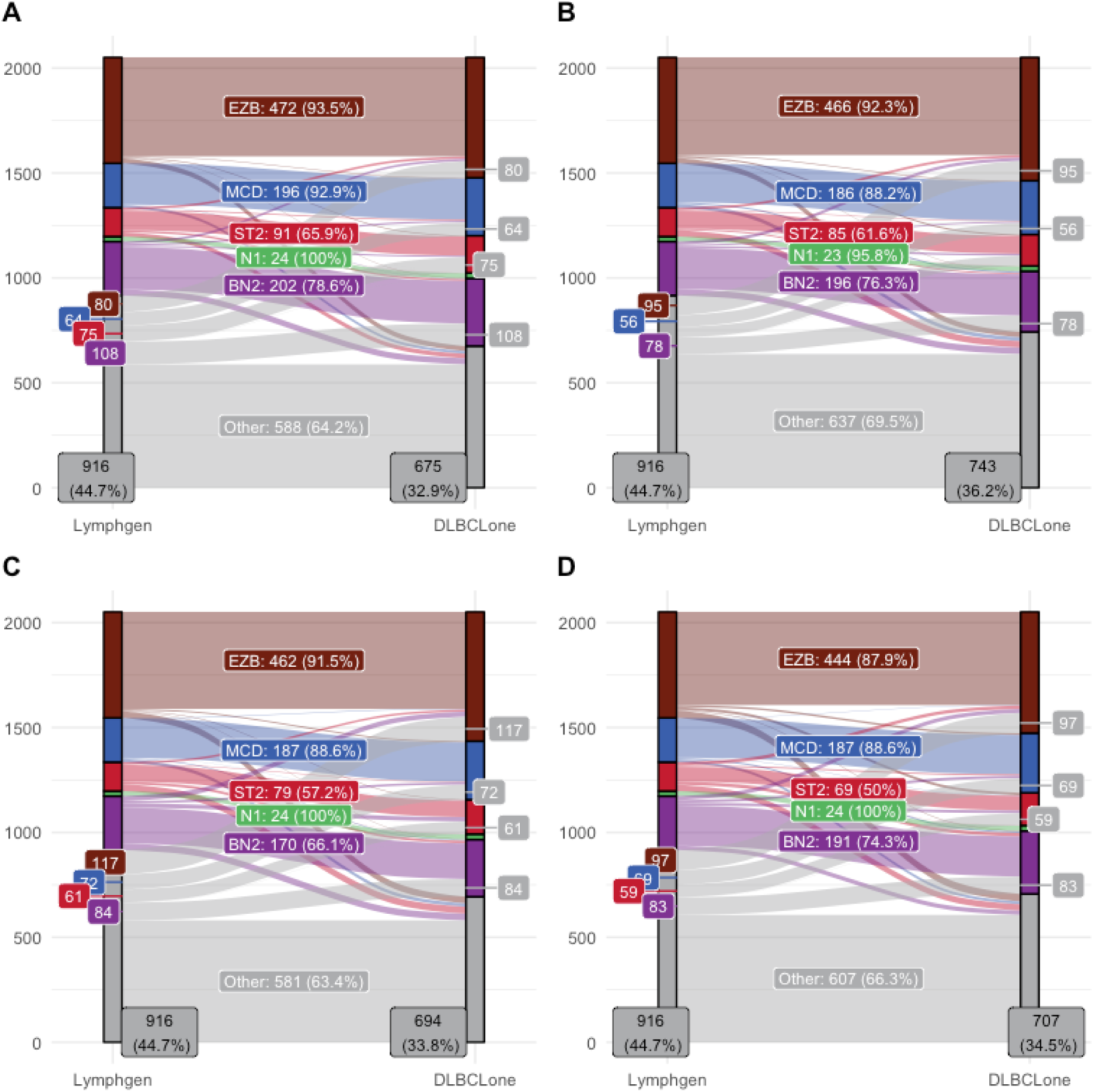
The training accuracy and classification results are summarized for DLBCLone models trained with four different feature sets **A)** Genes significantly associated with LymphGen from our post hoc analysis (n=85). **B)** LySeqST genes (n=125) **C)** Lacy genes (n=78) **D)** LymphPlex genes (n=37).

For a qualitative assessment, we visualized the classifications from our two best models alongside each sample’s genetic features(**Figure 6**). Samples are arranged to group those with discrepant assignments between LymphGen and DLBCLone. The samples reassigned from a LymphGen class to Other by DLBCLone represent a minority. Encouragingly, while officially unclassified, the greedy DLBCLone assignment was correct for the majority of these. Their assignment to Other by our optimized approach likely reflects different factors considered by LymphGen for in/out membership. In contrast, we were intrigued by the substantial number of samples that transitioned from LymphGen-Other to a class. Considering the number of samples reassigned to each class, the we saw the most growth of BN2, followed by ST2, MCD and EZB. In theory, these result from class-informative features in the full gene set that affect DLBCLone decisions but are considered less important (or ignored) by LymphGen. Indeed, LymphGen considers mutations in only 64 genes of the 125 covered by the LySeqST panel and the 85 features identified from our *post hoc* analysis.

**Figure 6:**
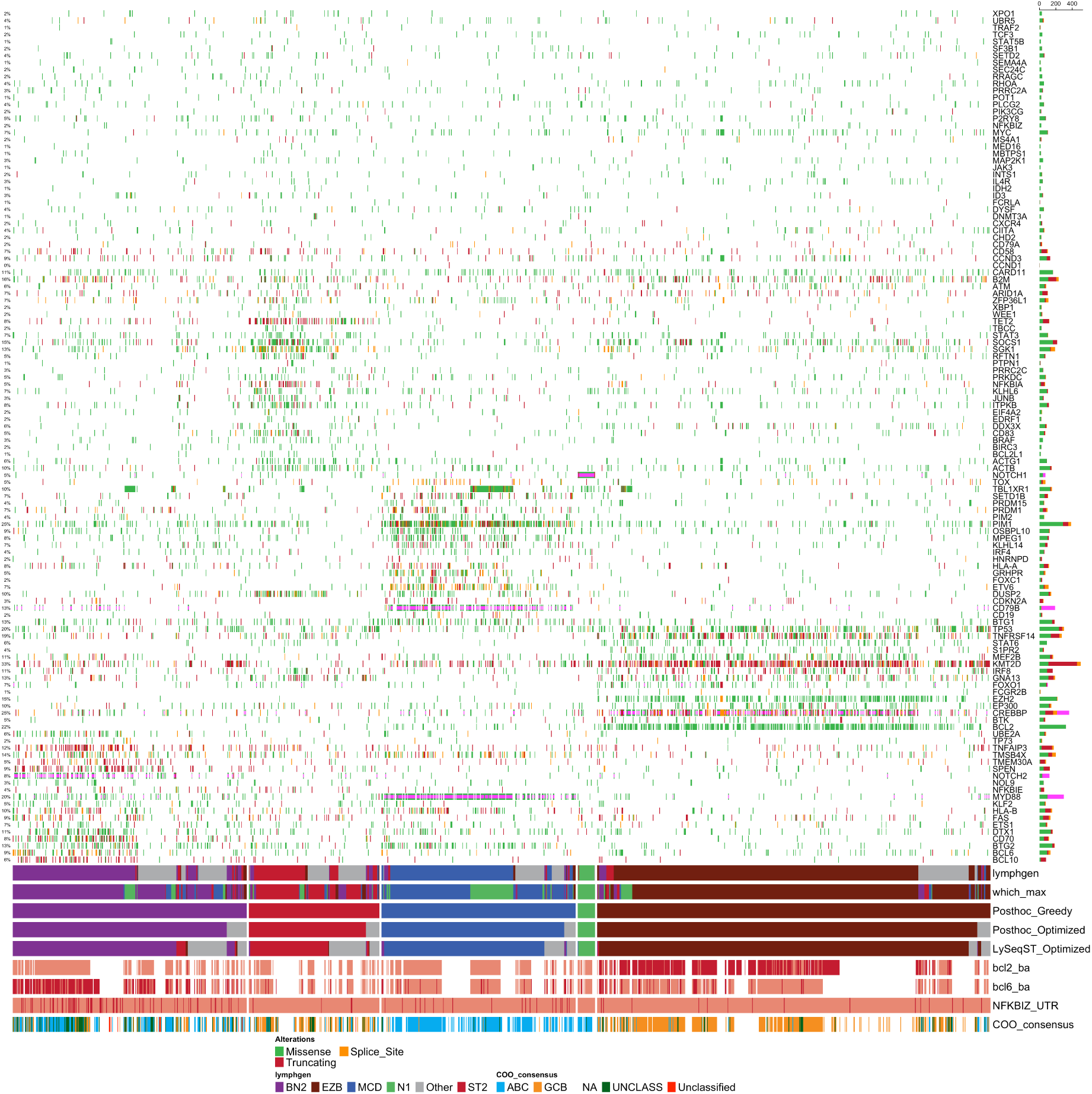
Mutation patterns of DLBCLs classified by the model trained with LySeqST. This oncoplot includes all samples from the training cohort except those classified as Other by both LymphGen and DLBCLone. The genes are arranged from bottom to top based on the association between their mutations and BN2, EZB, MCD, N1 or ST2 (respectively). The remaining genes (top) were not significantly associated with any LymphGen class. Each sample is annotated at the bottom with its original LymphGen class along with the DLBCLone greedy and optimized predictions using the classifier derived from the LymphGen-enriched features identified through our post hoc analysis of LymphGen (Figure 5A). DLBCLone assignments from the classifier trained using the LySeqST panel are also shown (Figure 5B). For comparison, the *which_max* label is a naive class inferred by taking the maximal count of classenriched mutations.

### Validation of classifiers on external data

We assessed generalizability using two external datasets. The first was a 1,001-patient cohort, of whom 337 underwent exome sequencing that the authors used to assign LymphGen labels (the Ruijin cohort)^12^. We applied the DLBCLone model trained on LymphPlex features to infer LymphGen labels for all cases. On the exome subset, our classification rate was 51.2%, exceeding LymphGen’s 36.4% (**Figure 7A**), with an overall accuracy of 0.70. Two caveats apply. First, mutations in genes not on the LymphPlex panel were available to LymphGen but hidden from DLBCLone; notably, 123/1,001 samples had zero LymphPlex genes mutated, forcing an “Other” assignment. Second, the LymphGen results include the A53 class, which DLBCLone was not trained to predict, so the nine A53 samples are counted as errors. Excluding A53, accuracies were similar for DLBCLone and LymphGen (0.728 vs 0.751), and DLBCLone achieved a slightly higher classification rate (52.4% vs 49.5%).

**Figure 7:**
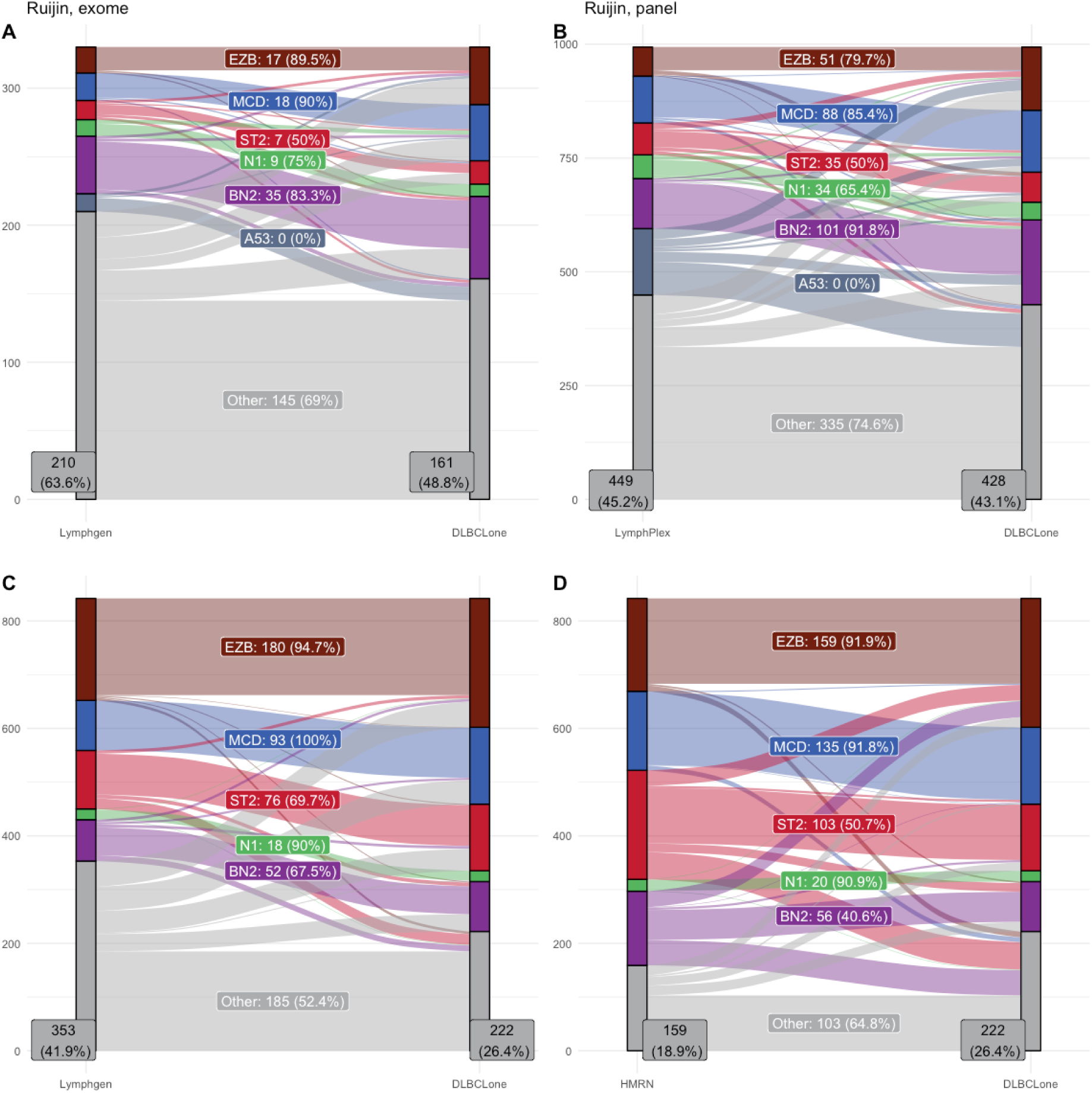
Performance of DLBCLone models trained on external data. **A)** Concordance between LymphGen class labels from the Ruijin exome cohort and DLBCLone approximations of LymphGen using the same feature set. **B)** Concordance between LymphPlex and DLBCLone approximations of LymphGen using the LymphPlex panel data from the Ruijin cohort. **C)** Correspondence between LymphGen class labels from the Lacy study and DLBCLone approximations of LymphGen using the same feature set. **D)** Correspondence between HMRN class labels from the Lacy cohort and DLBCLone approximations of LymphGen using the same feature set. ST2 was conidered equivalent to both SOCS1/SGK1 and TET2/SGK1.

The remainder of the Ruijin cohort was only subjected to panel-based sequencing along with FISH for BCL2 and BCL6 translocations and, together, these were used by the authors to infer LymphGen-like classes using their LymphPlex algorithm^12^. Comparing LymphPlex labels to DLBCLone’s optimized labels showed our ability to achieve a higher classification rate (55% vs 74.3%)(**Figure 7B**). If we consider LymphPlex as truth, the accuracy of DLBCLone assignments was 0.648. Again, all samples assigned A53 by LymphPlex were, by definition, considered mis-classified by DLBCLone. This metric also considers the 114 LymphPlex-Other samples given a different class by DLBClone. At least some of the discrepancies represent situations in which DLBCLone outperforms LymphPlex.

We trained another classifier for another in-house panel, which was used to sequence a cohort of 323 patients (the BC cohort)^16^. Remarkably, on the validation cohort, DLBCLone (optimized) achieved a classification rate of 88% with an accuracy of 0.85 (excluding LymphGen-Other and A53). An additional 98 samples that were either Other or A53 were assigned to one of the available classes (26 BN2, 36 EZB, 21 MCD, 2 N1 and 13 ST2). The mutation profiles of these re-classified samples, in general, show patterns of translocations, mutations and gene expression (COO) subgroup consistent with their DLBCLone-assigned class (**Figure 8**; **Supplemental Table S2**). For example, a substantial number of BCL6-translocated samples were reclassified from Other to BN2 by DLBCLone and several ABC samples with the *MYD88* hot spot mutation were added to MCD. Similarly, the majority of the new EZB and ST2 samples were GCB. Importantly, these molecular features were intentionally hidden during the training of this classifer to be consistent with the data used for LymphGen assignment in Wright *et al*^20^.

**Figure 8:**
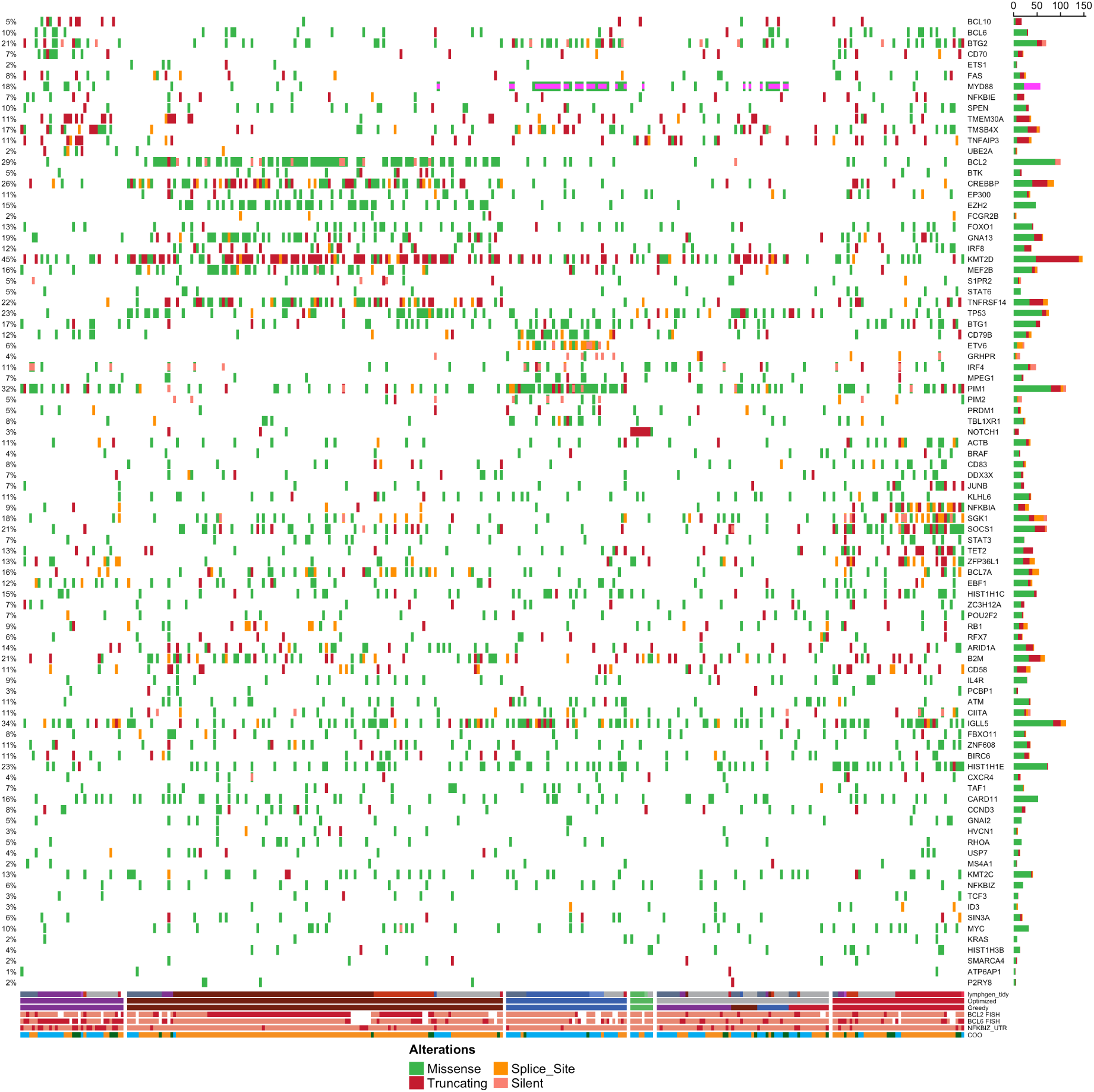
Mutation patterns of DLBCLs from BC validation cohort. This oncoplot includes all samples that were subjected to targeted sequencing in our previous study^16^. These samples were also used to validate the LymphGen algorithm^5^. The genes are arranged from bottom to top based on the association between their mutations and BN2, EZB, MCD, N1 or ST2 (respectively). The remaining genes (bottom) were not significantly associated with any LymphGen class. Each sample is annotated at the bottom with its LymphGen class from Wright *et al*. along with the DLBCLone greedy and optimized predictions using a classifier trained on the same features in Figure 2D, which included mutation status of the *NFKBIZ* 3 UTR. *BCL2* and *BCL6* translocation status is shown but this information was hidden during DLB-CLone training.

**Figure 9:**
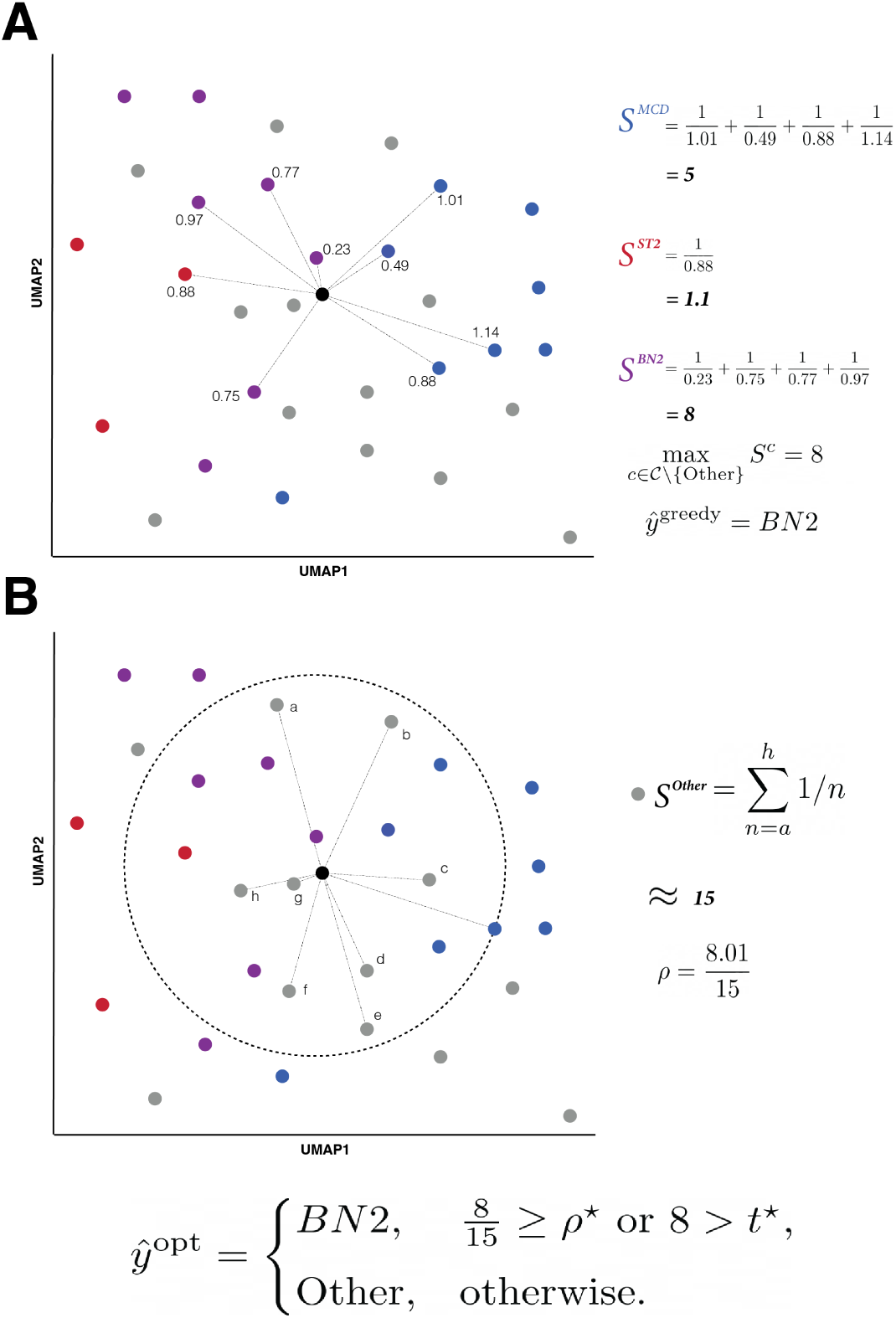
Overview of the DLBCLone procedure for inferring the class for a given sample after projection onto the UMAP alongside the training data for a given K (K = 9). **A)** The score for each class is first calculated using the Euclidean distances to the nearest K non-Other samples. The greedy label is the class with the maximal value. **B)** Next, we calculate the ratio between the maximal class score and the local Other score (*ρ*), which is similarly derived from the distances to the Other samples in the same radius. The optimized class assignment retains the greedy label if the magnitude of the original class score exceeds the threshold *τ*^⋆^ or, failing that, *ρ* exceeds a second threshold *ρ*^⋆^.

## Discussion

Real-world applications of genetic subtyping for DLBCL are only beginning to emerge, commonly making use of targeted sequencing rather than exomes or genomes^14,21–23^. As expected, performance of classifiers trained on comprehensive data degrades as the available feature space shrinks. In silico, a ∼400-gene panel classified 58% of the original cohort from Schmitz et al. with ∼92% overall accuracy when CNVs were included^4^, while a real-world application reached ∼55%. Another study using a ∼300-gene custom panel performed similarly (∼53%)^8,21^. In practice, nearly half of DLBCL tumours sequenced with these panels are left without a subtype assignment. We found a substantial number of genes whose mutation status is strongly associated with a LymphGen class (**Table 1**), highlighting a broader feature space that could enhance classification rates and inform custom panel design for DLBCL.

Notably, some of the genetic features we identified are already leveraged by DLBClass, possibly contributing to its higher classification rate. Nonetheless, with exome data, DLBClass assigns ∼75% with high confidence, leaving the remainder effectively unclassified. Limiting to mutations from the Lacy panel further reduced this to 61% high-confidence assignments^6,8,10^. This underscores the need for an approach that can achieve high classification rates with complete genetic data and maintain them within the constraints of a given panel. By training directly on the features queried by a panel, we consistently achieved higher classification rates on the same inputs while maintaining competitive per-class accuracy.

We focused on LymphGen in this study because DLBClass requires high-quality copy-number profiles that are harder to resolve from many clinical panels^10^. In principle, however, DLBCLone can be used to emulate DLBClass because the framework is feature-agnostic and can incorporate copy-number states and other events (e.g., SV/FISH) as additional features. Orthogonal molecular descriptors can also be included as categorical inputs. We did not explore CNV-augmented models here, but this is a natural extension that should improve recovery of CNV-driven classes (e.g., A53/C2) and help disambiguate overlap when mutation-only panels are sparse. Without robust CNV input, we anticipate reduced performance specifically for C2, mirroring DLBClass’s dependence on copy number.

While the performance of DLBCLone is encouraging, our results leave some questions unresolved. Some disagreements with LymphGen likely reflect true biological signal—features that are informative but under-weighted or unused by fixed classifiers—rather than error; the same logic extends to LymphGen-Other samples that join a true class. Since discrepancies exist between LymphGen and DLBClass, the true class boundaries remain unsettled across taxonomies. Meta-features improved recovery of N1, whereas other classes emerged more naturally from individual features. Based on our post hoc analysis of LymphGen-classified training samples, we identified TBL1XR1 as enriched for mutations in N1 (**Table 1**). TBL1XR1 mutations are also strongly associated with the MCD genetic subgroup and can promote the establishment of extranodal lymphomas^24^, motivating further work to determine whether NOTCH1-mutated cases form a distinct class or extend MCD.

In this evolving landscape, DLBCLone offers immediate, practical value. Without any new data, it can be used to evaluate the theoretical accuracy of a panel design. It is provided as an open-source R package and can be integrated into pipelines or wrapped in a Shiny user interface^25^. Our training workflow prioritizes flexibility, allowing users to tune the balance between classification rate and accuracy. Each trained model can be frozen and exported for local reuse or shared for broader application, with deterministic projection ensuring reproducible prospective assignment of samples. In short, DLBCLone provides a taxonomy-agnostic approach to DLBCL subtyping that may help bring subtype labeling within reach of more scientists, clinicians, and patients.

## Methods

### Patient samples

We assembled a cohort of 2542 patients diagnosed with diffuse large B-cell lymphoma (DLBCL). The raw sequencing data used in this study is available from the European Genome-phenome Archive at the European Bioinformatics Institute (EGAS00001007053, EGAS00001001709, EGAS00001004285, EGAS00001002936, EGAS50000000328, EGAS00001004469, EGAS00001006327, EGAS00001001600, EGAS00001001692, EGAS00001002606)^16,26–34^ and can be accessed through a request as detailed on the EGA website. In addition, the analysis was supplemented with the DLBCL whole-genome (WGS) and whole-exome (WES) data available from dbGAP (accessions phs000235, phs000328, phs000527, phs000450, and phs003023)^35–42^.

### Mutation analysis

All WGS and WES raw sequencing data was analyzed for sequencing quality according to the previously published guidelines^43^. Simple somatic mutations (SSM) were detected as previously described in detail^37^. Briefly, SSM were called using SLMS-3 pipeline that amalgamates Strelka^44^, SAGE, Lofreq^45^, and mutect^46^ to report somatic variants only supported by at least three algorithms. When the raw data was aligned to a different genome build, the identified SSM were converted to the GRCh37 genome build coordinates using Crossmap^47^. The detected variants were further annotated using command line vcf2maf (version 1.6.18) and Variant Effect Predictor (cache version 86).

When available, the fluorescence in situ hybridization (FISH) was performed for *BCL2*, *MYC*, and *BCL6*, and the FISH results were used for structural variant (SV) annotation. In addition, all WGS tumours were also analyzed for SV breakpoints using both Manta (version 1.6.0)^48^ and GRIDSS2.0 (version 2.9.4)^49^. SV calls from both tools were combined using the BioConductor StructuralVariantAnnotation package. Samples with SV connecting *MYC*, *BCL2*, or *BCL6* associated with a known recurrent partner locus were also considered positive.

### Post hoc inference of genetic features associated with LymphGen classes

We determined which individual genetic features were enriched in a LymphGen class using Fisher’s exact tests. The analysis included only training-cohort samples with unambiguous LymphGen assignments (i.e., excluding Other and composite labels). For every feature–class pair, we formed a 2×2 table comparing the number of mutated and non-mutated tumors inside the class versus outside the class, and performed a two-sided Fisher’s exact test. Unless noted otherwise, features were considered positively associated with a class when they met a nominal threshold (e.g., < 0.01) and showed enrichment in the index class (OR > 1). The resulting per-class feature lists were used to interpret neighborhood structure, guide meta-feature design, and visualize discrepant assignments in downstream analyses.

### Encoding and weighting genetic features

LymphGen uses a basic binary encoding to designate the presence and absence of any mutation in a gene or a copy number event (when provided). A notable restriction of this is that a synonymous mutation in a gene is given equal weight as a missense mutation. DLBClass employs a more complicated base-4 encoding, using 1 for silent mutations and low-level CNVs, 2 for non-silent mutations and high-level CNVs and 3 for SVs and 0 for the absence of a feature^10^.

Using this encoding, we found that UMAP tended to force samples with SVs away from the remaining samples. For our analyses used base-3 encoding except where this was not possible due to limitations of the data sets. Specifically, we reserved 1 for silent mutations in certain genes and 2 for either non-silent mutations or SV. We did not find reasonable justification to include silent mutations in every gene. Instead, we limit this to the genes known to be affected by aberrant somatic hypermutation (aSHM). As some data sets have already removed/hidden silent mutations, we also examined the effect of binary encoding. There was no obvious global differences in the layout of samples for the UMAP derived from these two alternative encodings (*not shown*).

### Latent representation for neighbor identification

We first build a reference embedding from the training cohort. Starting with the encoded feature matrix *X* (base-3 by default; binary where silent calls are unavailable), we optionally append meta-features and apply per-feature weights to obtain *X*^′^. We then fit UMAP on *X*^′^ with 2D output, using cosine distance, a fixed random seed, and stored transform. This yields a stable coordinate system (the “reference map”) that is retained for subsequent use in training and prediction.

For deployment and for cross-dataset comparability, we freeze the reference map and project samples with UMAP’s out-of-sample transform (e.g., uwot::umap_transform) using the same preprocessing and seed. This step *does not refit* the layout or move the reference points; each sample is placed near its training-set neighbors in the existing space. As a consequence, projected coordinates for training samples are not identical their original fit coordinates—the original layout reflects a global optimization over all points, whereas the transform places points relative to a fixed neighbor graph. Crucially, local neighborhoods are preserved, and the placement of new samples is deterministic and reproducible across runs and machines.

### Training

DLBCLone uses a K-nearest neighbors (KNN) approach to assign DLBCLs to any genetic classification system (**Figure 8**). LymphGen was used for this analysis because we were able to run their classifier on our samples. The algorithm relies on the local neighborhood of samples in the UMAP embedding. Importantly, because the UMAP algorithm is influenced by its input data, any subsequent addition of data would normally change the latent representation. We mitigate this by generating a model-based UMAP for a given set of features using a fixed seed and stable reference cohort. Samples are then projected using its out-of-sample transform, which positions each sample near its training-set neighbors while holding the original embedding fixed^50^. This step is accomplished by the make_and_annotate_umap function in our R package (GAMBLR.predict). At this stage, there is no risk of overfitting this model because only the mutations, not the class labels, are considered by UMAP.

The next phase in training is to assign every sample to a class (not Other) with the exception of those with zero features, which are assigned Other. The Euclidean distances *d* between a sample and all K-nearest neighbors in the training set is calculated from the 2-dimensional UMAP coordinates of all training samples. At this stage, the samples in the outgroup (Other) are not considered neigbors.

For a test sample *i*, we retrieve neighbours from the embedded training set, exclude Other neighbours when forming the non-Other neighbourhood, and add a small constant *ε* = 10^−1^ to distances (to avoid dividing by zero). Neighbour weights are inverse-distance:

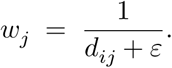

Let 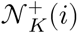 be the first *K* nearest non-Other neighbours, and 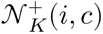 those among them labeled as class *c* in the ground truth. For each of the main classes *c* ≠ Other, we define the score for class *c*, *S*_*c*_(*i*) as

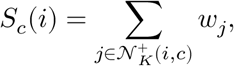

Using this score, we obtain the greedy DLBCLone label for sample *i*, *ŷ*^greedy^(*i*)

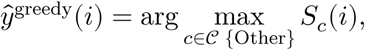

with ties resolved deterministically by a fixed class order. These computations are implemented in `weighted_knn_predict_with_conf` and `process_votes`.

We separately quantify the relative density of Other samples in the neighborhood.

Let 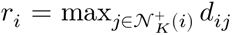 be the radius to the *K*^th^ non-Other neighbour. We define the Other score for sample *i S*_Other_ (*i*), as as the sum of the inverse of distances to Other neighbours *inside* this radius,

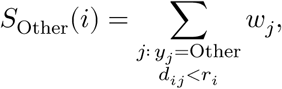

and the purity ratio

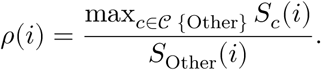

To limit over-assignment, we gate the greedy label using (i) the ratio *ρ*(*i*) and (ii) an absolute top score *S*_max_(*i*) = max_*c*_ *S*_*c*_(*i*). Thresholds (*ρ*^⋆^, *t*^⋆^) are selected by grid search to maximize macro (per-class) balanced accuracy (optionally with a cap on classification rate). The final prediction is

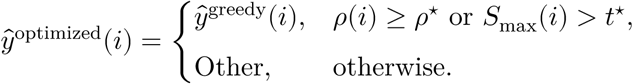

### Hyperparameter selection

We select *K* over an odd integer grid (here *K* ∈ {7, 9, …, 19}) and (*ρ*^⋆^, *t*^⋆^) by grid search on the training set to maximize a user-chosen objective. By default, the objective is macro balanced accuracy **excluding** ground-truth Other cases. Alternatives include overall accuracy or a weighted harmonic mean of balanced accuracy, classification rate, and concordance. Ties in arg max are broken by a fixed class order (user-configurable). This procedure is implemented in `DLBCLone_optimize_params`.

### Classification

For a given feature set, each sample *i* is represented by a base-3 vector *x*_*i*_ ∈ {0, 2}^*G*^ (gene- or event-level; e.g., *MYD88* hotspot, *BCL2*/*BCL6* translocation). Missing features are treated as absent (0). If synonymous mutations are unavailable, a binary encoding. If feature weights or meta-features were generated during training, the same treatment is applied to the new (previously unseen) samples. The frozen UMAP originally fit from the training data is restored (e.g. from disk). New samples are projected with UMAP transform using a fixed seed, and then assigned a greedy and optimized label via the KNN scores and the learned (*K*, *ρ*^⋆^, *t*^⋆^) thresholds (`DLBCLone_predict`).

## Data Availability

No new data was produced in this study. All data used in this study is available through the repositories and study accessions detailed in the text.

## Acknowledgements

The authors gratefully acknowledge funding support from the Canadian Institutes of Health Research. This study was also supported by a Program Project Grant from the Terry Fox Research Institute (#1061,#1108).

